# A Conceptual Approach to Comprehensive Interdisciplinary Cognitive Rehabilitation: A description using the TIDieR-Rehab Framework

**DOI:** 10.1101/2025.04.14.25325446

**Authors:** Lisa A.S. Walker, Marcia Finlayson, Julie Cameron, Sarah J. Donkers, Katherine Knox, Nancy Mayo, Michelle Ploughman, Kayleigh Canlas, Michaela Jacksch, Sharon Liu, Lara Pilutti MSCanRehab

## Abstract

Cognitive impairment is highly prevalent in people with progressive multiple sclerosis. Computerized cognitive training techniques have demonstrated some success at targeting core cognitive processes but are criticized for lack of transfer to real-world tasks. Research in physical rehabilitation has demonstrated the efficacy of high volume, moderate intensity task-specific training (TST) that is monitored, progressed and ideally practiced in real-world environments to enhance learning, transferability and support neurorecovery of motor functions. Findings demonstrate that TST influences activity and participation outcomes, leads to improved cortical reorganization, and focuses on activities that align with a person’s daily life and specific goals. Our MSCanRehab group adapted the same principles to develop a novel comprehensive interdisciplinary approach to cognitive rehabilitation that combines core cognitive process training (CCPT), TST and a modified Cognitive Orientation to Daily Occupational Performance (CO-OP) approach. The cognitive TST was individualized to participant goals identified using the Canadian Occupational Performance Measure and further tailored based on their areas of cognitive weakness. A modified CO-OP process was utilized that incorporated CCPT and cognitive TST personalized to meet meaningful participant goals and improve occupational performance. Cognitive TST utilized goal-directed practice and repetition focused on cognitive skills (vs. impairments) using real-world tasks. Task complexity and challenge was gradually increased and progressively adapted with emphasis on active participation and problem solving. A case example demonstrates how the intervention was implemented. Components of the intervention are described using the TIDieR-Rehab checklist to ensure a comprehensive description, with emphasis on essential elements and dosage parameters.

**Highlights:**

- Computerized cognitive training alone can lack generalizability to real-world settings
- The task-specific training approach used in physical rehabilitation can be adapted to address cognitive rehabilitation goals
- A modified Cognitive Orientation to Daily Occupational Performance (CO-OP) process that incorporates core cognitive processes training with cognitive task-specific training provides a personalized and adaptable approach to cognitive rehabilitation in people with progressive MS

## Introduction

Cognitive impairment is prevalent in people with multiple sclerosis (MS) with rates of impairment higher in those with progressive forms. Up to 79% of people with secondary progressive MS and 91% of those with primary progressive MS experience cognitive impairment(1). Deficits lead to functional limitations that impede effective engagement in meaningful daily tasks(2, 3). In turn, functional limitations can negatively impact mood, with depression symptoms further restricting the ability of people living with MS to pursue leisure and outdoor activities(4). Given their dissatisfaction due to lack of access to treatments focusing on participation in cognitively-based daily occupations, there is a need to pursue research addressing this gap(5).

This article describes a comprehensive interdisciplinary approach to cognitive rehabilitation that combines core cognitive process training (CCPT), principles of cognitive task-specific training, and methods associated with a modified Cognitive Orientation to Daily Occupational Performance (CO-OP) approach (described below). It is important to begin with definitions. The American Congress of Rehabilitation Medicine defines *cognitive rehabilitation* “as a systematic, functionally oriented service of therapeutic cognitive activities, based on an assessment and understanding of the person’s brain-behavioural deficits” (6)(p. 63). The goal of cognitive rehabilitation is to achieve functional improvement utilizing restorative strategies to facilitate neuroplasticity and/or compensatory strategies(6). *Cognitive training* can be considered an element of cognitive rehabilitation. For the current purposes, we reference CCPT in which a specific aspect of cognition (e.g., divided attention, processing speed, etc.) is targeted for restoration utilizing computer software. *Cognitive stimulation* refers to incorporating progressively complex cognitively stimulating activities (here in the context of TST described below) that simulate the demands of a real-world context.

There has been much focus on computerized cognitive training methods to target *core cognitive processes* and these have demonstrated some success(7), with at least one study of persons with progressive MS reporting benefits in learning, memory and processing speed (8). However, major criticisms include lack of generalizability, lack of direct link to the person’s goals, and lack of transfer of benefits to daily activities and untrained tasks, as well as whether treatment meaningfully impacts daily functioning(9, 10). MS rehabilitation should be patient-centered, focused on individuals’ complex needs, and performed by a multidisciplinary team(11). Rather than focusing on symptoms, the primary emphasis should be on the goal of optimizing engagement in daily life(12).

When people with MS undergo physical rehabilitation, *task-specific training* (TST) is one approach to support recovery of motor functions. TST influences activity and participation outcomes more than simply targeting a specific motor impairment(13) and leads to improved cortical reorganization(14). TST focuses on practicing functionally relevant tasks that align with daily life and specific goals using repetitive and targeted practice of tasks that mimic real-world activities. It is grounded in principles of learning and neuroplasticity (15, 16). Task complexity should increase with time to maintain an optimal level of challenge(17). Challenge can be manipulated by adjusting task parameters (e.g., number of steps), environmental/contextual factors (e.g., dual-tasking), and personal factors (e.g., fatigue level). Feedback is an important part of skill acquisition(16), which might be intrinsic (e.g., cognitive effort required), or extrinsic (e.g., verbal cues). Last, training takes place in an environmental context closely resembling natural/real-world contexts in which tasks are performed to promote generalization of functional gains beyond clinical settings. Motor TST is evidence-based and effective in training upper limb functions and gait rehabilitation(18). While motor TST has received much attention(18), there is a paucity of research on cognitive TST(19), with only one recent study in MS(20).

The principles of motor TST can apply to *cognitive TST*(19). The training program can be individualized and focus on improving performance of context-specific cognitive tasks through goal-directed practice and repetition focused on cognitive skills (vs. impairments). Functional task domains (e.g., household responsibilities, shopping, workplace-related, etc.) should be selected based on participant’s goals and should, to the extent possible, be directly related to goals (e.g., if the person selected banking as a goal, then activities related to financial management should be used). Since cognitive function emerges from interactions of the task, individual, and environment, task components and environmental constraints should be manipulated to mimic performance in real-world settings(21). Training parameters are guided by principles of cognitive learning and experience-dependent neuroplasticity(14). Intensity should be monitored over time, preferably objectively, and as a participant makes gains/improvements, tasks should progress to maintain optimal challenge level.

The CO-OP (Cognitive Orientation to daily Occupational Performance) approach, a patient-centred method employing collaborative goal-setting, dynamic performance analysis, cognitive strategy use, guided discovery, and enabling principles(22) was recently applied in a study of people with MS(20). The CO-OP approach led to improvements in daily activity performance and quality of life in a sample of 6 people with Expanded Disability Status Scale between 3.5 and 5 and a Montreal Cognitive Assessment score above 26/30. CO-OP involves individualized goal-setting, teaching global cognitive strategies to promote skill acquisition (goal-plan-do-check), domain-specific strategies to help overcome problems performing activities, and generalizing to other environments and transfer to other skills(22). Given that computerized cognitive rehabilitation techniques have proven successful at improving core cognitive processes in those with progressive MS(23), and motor TST has demonstrated benefits in MS(24), the MSCanRehab research team combined these (described in detail below) and adapted the CO-OP framework to generate an innovative interdisciplinary approach to cognitive rehabilitation. This more comprehensive approach includes CCPT, informed by objective measurement, with patient-driven and goal-directed TST.

The current paper describes our approach, which is a component of a larger interventional study investigating the feasibility of brain priming paired with comprehensive cognitive and motor rehabilitation to restore cognitive or motor abilities in people with progressive MS. In terms of the cognitive rehabilitation component of the intervention relevant to the current paper, CCPT (using RehaCom(25)) and TST are administered over 8 weeks guided by participant goals as identified through the Canadian Occupational Performance Measure (COPM)(26). Feasibility and preliminary efficacy of the larger intervention will be reported elsewhere.

### Aim

The overarching aim of the current paper is to describe the comprehensive interdisciplinary approach to cognitive rehabilitation that combines CCPT, principles of TST and methods associated with a modified CO-OP approach emphasizing essential elements.

## Methods

We used the Template for Intervention Description and Replication (TIDieR) checklist(27) including information specific to rehabilitation trials (TIDieR-Rehab checklist) to describe the intervention. (27-29) (see Appendix A). The TIDieR-Rehab checklist contents are presented in a different order than indicated in Appendix A to improve flow. Ethics approval was received from the University of Ottawa Research Ethics Board.

### Personnel

The study was developed and conducted by MSCanRehab; a multi-disciplinary group of Canadian MS rehabilitation researchers. Primary team members involved in the cognitive protocol were a licensed occupational therapist (MF) and a licensed clinical neuropsychologist (LW), who were responsible for this portion of study design and oversight, and a second occupational therapist who served as the interventionist (JC) and supervisor to the research assistants/therapists delivering the CCPT and cognitive TST. The interventionist and the therapists/research assistants were trained by MF and LW in the study protocol, Canadian Occupational Performance Measure (COPM) (used for goal setting), RehaCom (used for CCPT), and the cognitive TST manual. Interventionists and therapists/research assistants also completed COPM on-line training(30). JC designed each participant’s personalized intervention in consultation with MF and LW.

### Setting

Cognitive rehabilitation was delivered in a university-based research laboratory. All cognitive rehabilitation procedures took place in-person, with participants working with one or more interventionists/research assistants depending on TST complexity. CCPT (see below) took place in a quiet distraction-free laboratory room. A research assistant was present to set up the software and ensure the participant could manage the interface. Thereafter, they remained available but out of sight to reduce distraction. For the TST, the lab environment included access to various home-like settings (e.g., laundry facilities, kitchen area). This ensured tasks were completed in an environment as close to real-world as possible. An important consideration is to ensure lab environments can include resources specific to participant goals.

### Participants

Participants for the cognitive rehabilitation aspect of the intervention were adults (>18 years of age) living with progressive MS (or living with MS for ≥10 years) who identified self-reported cognitive challenges that interfered with functioning in daily life and were willing to complete testing and training visits. Exclusion criteria involved the diagnosis of another neurodegenerative condition, traumatic brain injury, learning disability, attention-deficit/hyperactivity disorder (ADHD), or other major psychiatric disorder. Other exclusion criteria related to the larger study involved the inability to undergo non-invasive brain stimulation; or the presence of uncontrolled cardiovascular, metabolic, renal, or other medical conditions that may impact exercise participation. Four participants met criteria and identified cognitive goals for occupational performance. All participants completed the comprehensive cognitive rehabilitation component of the intervention.

### Intervention Support Materials

#### Cognitive Task-specific Training (TST) Manual

A training manual was developed by LW, MF and SD for interventionists/research assistants to ensure intervention consistency. The manual included team contact information, overview of the larger study, intervention overview with theoretical background/rationale, and procedural information (e.g., team huddle, intervention session details, instructions for increasing task complexity).

#### Canadian Occupational Performance Measure (COPM)

A widely used tool by occupational therapists to assist in identification of issues important to clients(26), the COPM is individualized and client-centered. The COPM captures an individual’s self-perception of performance and satisfaction with performance in daily life and is validated in MS(31). Issues identified are the basis of goals guiding the selection and progression of tasks for cognitive TST. The COPM was administered by the interventionist (JC) prior to the intervention.

#### Core Cognitive Process Training (CCPT)

RehaCom is computer-assisted cognitive training software that includes screening and training modules targeting *core cognitive processes* including: attention, memory, executive functions, visual field training and visuo-motor abilities (Hasomed, Magdeburg, Germany)(25). RehaCom is widely used in clinical rehabilitation settings, validated in MS(7), and utilized as a cognitive training tool in a large-scale randomized controlled trial in MS(23).

#### Cognitive TST Examples

Although intervention tasks were individualized and dependent upon participants’ stated goals, an Appendix to the Cognitive TST Manual outlined several potential goals a participant may identify and the TST tasks the interventionist could utilize to address and progress these goals over time. Examples included basic tasks with which to begin and then gradually increased task complexity by adjusting activity-specific factors or task parameters, environmental/contextual factors (e.g., introducing variations or distractors), incorporating dual-tasking, and progressing to simulating complications that may occur with task completion during real-life. For example, if a participant identified grocery shopping as a goal the basic task might be “make a weekly menu”. Building on this, a mildly challenging task might be “check what ingredients you have and make your shopping list for missing items”. A more moderately challenging task might be “identify what stores to visit based on list items”. Finally, a maximally challenging task might be “organize your shopping trip to visit all stores based on aisles/sections in each store”.

#### Goal-Plan-Predict-Do-Review Worksheet

The CO-OP approach utilizes a Goal-Plan-Do-Check approach, previously utilized in MS(20). This approach was modified for the current protocol to incorporate a greater emphasis on problem-solving to facilitate independence and generalizability to other daily tasks (Figure 1). *Goals* were identified by participants via the COPM. With interventionist guidance, participants were encouraged to *plan* what to do to achieve their goal, first by considering needed materials, and then by thinking about required steps to complete the activity. Participants were encouraged to *predict* what steps they may find challenging and what to do to make success more likely. While this step could be considered part of the planning process (as per the initial CO-OP approach), by making the prediction step more explicit, it encourages participants to independently problem-solve how to overcome anticipated challenges and emphasizes executive demands needed to effectively manage everyday tasks. Participants then *do* the task according to the steps they established. Finally, the interventionist helped them to *review* what worked, what did not work, and what they could do differently on the next attempt. This review step involves greater active effort than simply checking on whether a goal was met, instead requiring a more intentional and reflective evaluation. These modifications to the original CO-OP approach place a greater emphasis on executive functions given the required prediction (i.e., anticipation) and review (i.e., self-assessment).

**Figure 1.**
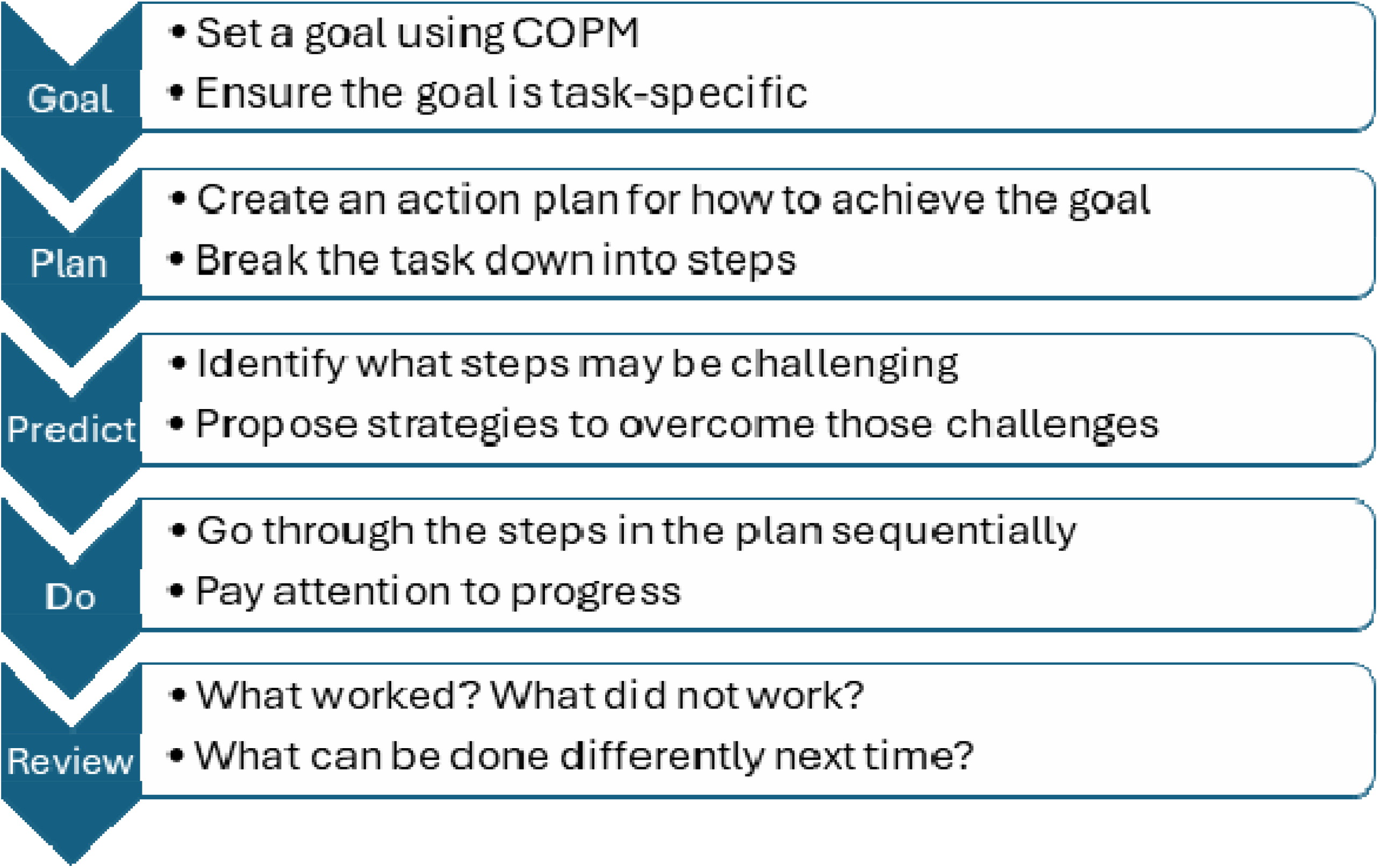
Goal-Plan-Predict-Do-Review.

#### Cognitive Training Tracking Sheet

For each cognitive training session, the research assistant completed a tracking sheet included as an appendix in the Cognitive TST manual. Relevant details of the CCPT (RehaCom) were recorded (e.g., start/end time, module(s) trained, adverse events). Similarly, details of TST were recorded (e.g., start/end time, task complexity, attempt number, confidence rating, task description, errors identified, adverse events). The tracking sheet helped ensure task fidelity.

### Intervention Process

The cognitive rehabilitation intervention is part of a larger study. Figure 2 depicts this aspect of the intervention process. Feasibility and preliminary efficacy outcomes for the intervention overall will be reported in future manuscripts. Times for essential elements of the intervention listed below are estimates based on planned session duration.

**Figure 2.**
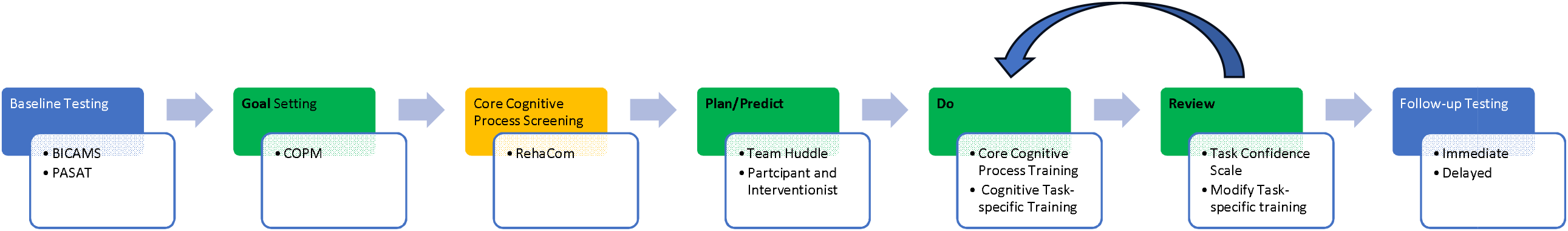
Process Diagram of Comprehensive Interdisciplinary Approach to Cognitive Rehabilitation. Legend: Blue boxes indicate the outcome measure testing, green boxes indicate steps in the Modified CO-OP component of the intervention which includes both core cognitive process training and cognitive task-specific training, the orange box indicates the core cognitive process screening, and bolded text represents the steps of the Goal-Plan-Predict-Do-Review worksheet.

#### Baseline Testing

Following enrollment, participants underwent baseline cognitive testing as a component of a larger battery, including the Brief International Cognitive Assessment for MS and Paced Auditory Serial Addition Test, conducted by research assistants in a session lasting approximately 20 minutes.

#### Goal Setting

Participants then completed an intake meeting with the interventionist who administered the COPM, lasting approximately 45 minutes. Their top two or three goals were identified.

#### Core Cognitive Process Screening

All participants completed five RehaCom screening modules: Alertness, Divided Attention, Working Memory, Memory for Words, and Logical Reasoning. This ensured that core cognitive processes most impacted in individuals with MS (i.e., processing speed, working memory, learning/memory, and executive functioning)(32), and most likely to relate to participant goals, were evaluated. Administration time was 20-30 minutes on average.

#### Plan/Predict

Following baseline assessments, team huddles of approximately 30 minutes, took place with the interventionist, OT, neuropsychologist and principal investigator. The interventionist shared participant goals and performance on RehaCom screening modules. RehaCom training modules were selected based on areas of cognitive weakness identified and alignment with key component cognitive processes underlying identified goals. The remainder of the huddle was devoted to TST planning. Functional⍰task⍰domains were selected based on participant’s goals, and to the extent possible, were directly related to those goals (e.g., if doing laundry was a goal, then laundry activities were used in the TST).

Following the huddle, the interventionist met with the participant and together devised a plan based on the Goal-Plan-Predict-Do-Review worksheet. The participant was encouraged to predict which aspects of the task may be challenging and propose strategies to overcome those challenges. This initial planning took approximately 15 minutes.

#### Do

Participants completed the CCPT at the beginning of each intervention session. This was immediately followed by cognitive TST individualized for each participant and focused on improving performance of context-specific cognitive⍰tasks⍰through goal-directed practice and repetition of cognitive skills practice (vs. impairments). The interventionist was present to ensure any person-specific modifications were incorporated as-needed (e.g., accommodate physical limitations). Each goal began with task training at a basic level and then progressed to mildly challenging, moderately challenging, and maximally challenging tasks. The⍰cognitively stimulating tasks were progressively adapted and emphasized active participation and problem solving. Task⍰components and environmental constraints were manipulated to mimic performance in real-world settings.

#### Review

After each task attempt, the participant, with interventionist assistance, conducted a progress review using the Task Confidence Scale (Figure 3) to help determine appropriate TST intensity. Once a participant rated themselves as moderately to extremely confident (i.e., scale rating of 6 to 10) in their ability to complete the task then difficulty was increased. Similarly, if after trying to execute the plan, the participant was not confident (e.g., 1 to 3 on the scale), then difficulty was reduced. As a participant made gains/improvements,⍰tasks were modified to maintain challenge level. Progress in TST was monitored using a Cognitive Training Tracking Sheet. The number of task repetitions completed varied depending on the person, as well as the nature and complexity of the task (on average, a task was repeated approximately 3 times before increasing complexity).

**Figure 3.**
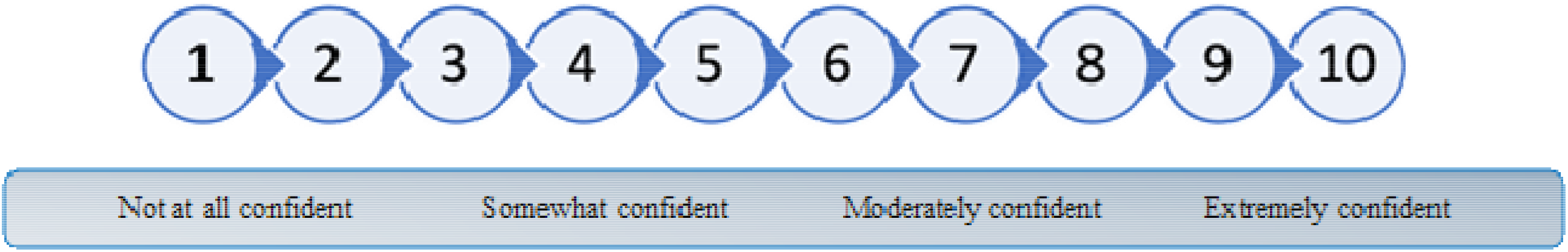
Task Confidence Scale.

#### Session Duration and Essential Elements Amount

The intervention took place over 8 weeks, with three sessions per week. To account for scheduling challenges and missed sessions, a buffer of 2 additional weeks was scheduled. Total planned intervention time was 40 minutes per session. Proportion of time allocated to CCPT and TST shifted over the course of the sessions. During the first week (i.e., three sessions), 40 minutes was allocated to CCPT (using RehaCom) alone to target core cognitive processes. Sessions in week 2 through 6 allocated 20 minutes each to CCPT and TST. Finally, in weeks 7 and 8, 10 minutes was allocated to CCPT and 30 minutes to TST. Increasing emphasis on TST was to empower participants to actively engage in meaningful activities, promote skill acquisition, and facilitate functional independence and participation in daily life.

#### Follow-up Testing

Within 1-2 weeks after intervention completion, participants underwent follow-up cognitive testing (RehaCom screening modules, Brief International Cognitive Assessment for MS and Paced Auditory Serial Addition Test). This was repeated 8-weeks post-intervention.

### Intervention Fidelity

A number of procedures were in place to ensure intervention fidelity. As noted above, huddles took place prior to intervention initiation for each participant. A clear plan was established, ensuring the intervention was individually tailored. In addition, all research assistants/therapists received unified training. Finally, regular check-ins with the interventionist and principal investigator were completed to ensure the intervention was delivered according to protocol. Protocol deviations and adverse events were recorded. Fidelity outcomes will be reported elsewhere once the larger brain priming and TST intervention feasibility study is complete.

### Case Example

This example illustrates the intervention for a middle-aged male with progressive MS. He first completed RehaCom screening, including assessments of Alertness without sound warning (Z Score = −0.01); Alertness with sound warning (−0.31); Divided Attention auditory modality (−2.38); Divided Attention visual modality (0.57); Working Memory, (1.71); Memory for Words (1.49); and Logical Reasoning (−0.05). Based on screening, Divided Attention cognitive training module was recommended.

The interventionist administered the COPM to identify occupational performance problems that affected his function in everyday life. The participant evaluated each occupational performance problem based on Importance (I), Performance (P), and Satisfaction (S) using a 10-point Likert scale (low scores indicate low Importance/Performance/Satisfaction). Four occupational performance problems were identified: (1) Remembering people’s names (I= 8, P =4, S= 2); (2) Recalling words in conversation (I= 10, P =6, S= 2); (3) Remembering podcast content (I= 9, P =7, S= 3); and (4) Prioritizing/planning daily activities (I= 8, P =2, S= 2).

The interventionist, OT, neuropsychologist and principal investigator then met for a team huddle to review occupational performance problems, RehaCom scores, and discuss TST ideas. The identified occupational performance problems, COPM scores and RehaCom Z-scores informed the TST activities. It was determined that three occupational performance problems (remembering people’ names; recalling words in conversation; and remembering podcast content) could be targeted in one functional task involving listening to audio clips from various podcasts. The participant’s fourth occupational performance problem (prioritizing/planning daily activities) was targeted in a series of TST sessions including education on planning/prioritizing strategies, goal setting and scheduling. Prior to TST sessions, the participant worked collaboratively with the interventionist to complete a Goal-Plan-Predict-Do-Review worksheet to plan steps needed to achieve his goals. The interventionist developed TST activities for each intervention week that were facilitated by the intervention team.

At each session, the participant completed the “Divided Attention” training module on RehaCom (10-40 minutes/session) prior to TST activities (20-40 minutes/session). Progression of TST difficulty level (basic, mild, moderate, maximum) was determined by the participant’s subjective feedback, objective performance on TST tasks, and confidence rating scores collected at the end of each session. Various factors, including task length, task novelty, and environmental distractions were modified over the course of the intervention to ensure tasks provided appropriate cognitive challenge. Table 1 includes descriptions of each task and how difficulty was adjusted.

### Future Directions

Research should be informed by those with lived experience to ensure more collaborative healthcare services that meet diverse needs of people with MS(33) and take into account experiences of those living with a complex disease, personal values, and different ways of knowing(33). Future work will incorporate feedback received from people living with MS enrolled in our feasibility study. Given our goal is to develop an intervention reflective of skills needed to engage in meaningful daily activities, easily implementable in a home setting, and tailored to person-specific needs of people living with MS, our group will consider this feedback moving forward. Decisions regarding a larger RCT will be made once feasibility and preliminary effects data are analysed.

### Conclusions

Our MSCanRehab group is the first to utilize a comprehensive interdisciplinary approach to cognitive rehabilitation that incorporates both CCPT and cognitive TST personalized to meet meaningful participant goals and improve occupational performance. Intervention components were described using the TIDieR-Rehab checklist with emphasis on essential elements and dosage parameters. Future publications will report intervention outcomes and results of the feasibility trial will inform a multi-centre efficacy trial.

## Data Availability

All data produced in the present study are available upon reasonable request to the authors.

## Acknowledgements

The authors wish to thank the people with MS who kindly participated in this study.

## CRediT Author Statement

**LW:** Conceptualization, Methodology, Writing – Original Draft, Visualization, Supervision; **MF:** Conceptualization, Methodology, Writing – Original Draft, Visualization, Supervision; **JC:** Methodology, Investigation, Data Curation, Writing – Original Draft, Supervision; **SJD:** Conceptualization, Methodology, Resources, Writing – Review & Editing, Supervision, Project Administration; **KK:** Conceptualization, Methodology, Writing – Review & Editing; **NM:** Conceptualization, Methodology, Data Curation, Writing – Review & Editing; **MP:** Conceptualization, Methodology, Resources, Writing – Review & Editing, Supervision; **KC:** Investigation, Data Curation; **MJ:** Investigation, Data Curation; **SL:** Investigation, Data Curation; **LP:** Conceptualization, Methodology, Validation, Investigation, Resources, Data Curation, Writing – Review & Editing; Supervision, Project Administration, Funding Acquisition.

## Appendix A. The TIDieR-Rehab checklist

| Item # | Item | Where located in primary paper | Other† (details) |
| --- | --- | --- | --- |
| **SECTION 1. BRIEF NAME** |  |  |  |
| 1 | Provide the name or a phrase that describes the intervention. | Title and Aim |  |
| **SECTION 2. WHY** |  |  |  |
| #2 | Describe any rationale, theory or goal of the elements essential to the intervention.<br>*Essential elements, also known as ‘active ingredients’, are the core components of the intervention that are expected to be linked to effects or outcomes of interest.* | Introduction |  |
| **SECTION 3. WHO** |  |  |  |
| *3 | Describe who the intervention is intended for. | Participants |  |
| **SECTION 4. WHEN** |  |  |  |
| *4 | Describe when the intervention commenced in relation to the onset or stage of the condition and/or other relevant events. | Participants |  |
| **SECTION 5. WHAT** |  |  |  |
| 5A | Materials: Describe any physical or informational materials used in the intervention, including those provided to participants or used in intervention delivery or in training of intervention providers. Provide information on where the materials can be accessed (for example, online appendix, URL). | Intervention Support Materials | Figures 1, 3 |
| 5B | Procedures: Describe each of the procedures, activities and/or processes used in the intervention, including any enabling or support activities. | Intervention Process | Figure 2 |
| **SECTION 6. WHO PROVIDED** |  |  |  |
| 6 | For each category of intervention provider (for example, psychologist, nursing assistant), describe their expertise, background and any specific training given. | Personnel |  |
| **SECTION 7. HOW** |  |  |  |
| 7 | Describe the modes of delivery (such as face to face or by some other mechanism, such as internet or telephone) of the intervention and whether it was provided individually or in a group. | Setting |  |
| **SECTION 8. WHERE** |  |  |  |
| #8 | Describe the type(s) of environment(s) where the intervention occurred, including any necessary infrastructure or relevant features.<br>*Environments and their features may include the relevant physical, social, cultural, economic, political and/or systemic context(s) of the intervention.* | Setting |  |
| **SECTION 9. HOW MUCH** |  |  |  |
| *9A | Session(s) duration: Specify the planned session(s) duration of the intervention. | Intervention Process |  |
| *9B | Essential elements amount: Specify the planned session(s) duration and/or repetitions of the essential elements of the intervention.<br>*Essential elements amount refers to the time and/or repetitions, within the total duration of a single session, that is spent ‘actively’ participating in the core components of the intervention.* | Intervention Process |  |
| *9C | Frequency: Specify the planned frequency of the intervention. | Intervention Process |  |
| *9D | Intervention length: Specify the planned overall length of the intervention. | Intervention Process |  |
| **SECTION 10. HOW CHALLENGING** |  |  |  |
| *10 | Describe the approach(es) used to set and monitor the intervention/task challenge level.<br>*Challenge may include the nominal, functional or perceived level of difficulty, effort, physiological intensity or cognitive load of an intervention/task at a given time and may be evaluated using subjective or objective measures.* | Intervention Process | Figure 3 |
| **SECTION 11. REGRESSION/PROGRESSION** |  |  |  |
| *11 | Describe the planned regression and/or progression of dosage parameter(s), including when and how.<br>*Dosage parameters refer to the amount (Section 9. How much) and challenge (Section 10. How challenging) of the intervention.* | Intervention Process | Figure 3 |
| **SECTION 12. PERSONALISATION** |  |  |  |
| *12A | Needs: If supplementary strategies were planned to enable the delivery of the essential elements of the intervention in response to specific individual or group needs, then describe what, why, when and how.<br>*Supplementary strategies refer to intervention adjuncts (for example, physical assistance, verbal cueing, props) that must be used by some individuals or groups to facilitate effective participation in the essential elements.* | Intervention Process |  |
| *12B | Preferences: If the intervention was planned to be adapted for personal preferences, then describe what, why, when and how. | Intervention Process |  |
| **SECTION 13. PROTOCOL DEVIATIONS** |  |  |  |
| #13 | If there were deviations in the intervention protocol during the course of the study, describe the changes (what, why, when and how). | n/a |  |
| **SECTION 14. HOW WELL** |  |  |  |
| 14A | Plan: If intervention adherence or fidelity was assessed, describe how and by whom, and if any strategies were used to maintain or improve fidelity, describe them. | Intervention Fidelity |  |
| 14B | Actual: If intervention adherence or fidelity was assessed, describe the extent to which the intervention was delivered as planned. | To be discussed in future feasibility paper |  |
| **SECTION 15. HARMS** |  |  |  |
| *15A | Plan: Describe the monitoring of adverse consequences.<br>*Adverse consequences include any negative, undesired effects related to the intervention, including physical, mental, social and/or spiritual effects.* | Intervention Fidelity |  |
| *15B | Actual: Describe any adverse consequences, including the number, seriousness and relatedness to the intervention. | To be discussed in future feasibility paper |  |
| *, new item unique to the TiDieR-Rehab checklist.<br>#, adapted original TiDieR item for the TiDieR-Rehab checklist.<br>TiDieR, Template for Intervention Description and Replication. |  |  |  |

